# Psychiatric Voice Biomarkers: Methodological flaws in pediatric populations

**DOI:** 10.1101/2025.10.13.25337901

**Authors:** Hammza Jabbar Abd Sattar Hamoudi, Mon-Ju Wu, Marsal Sanches, Cesar A. Soutullo, Carolina Olmos, Leslie K. Taylor, Giovanna Zunta-Soares, Jair C. Soares, Benson Mwangi

## Abstract

**Introduction:** Psychiatric assessments rely on patient self-reports, clinician observations, and standardized scales, while objective technological tools are currently not reliable enough to be utilized in a clinical setting. Voice may be utilized as a biomarker in different scenarios, including differential diagnosis, assessing symptom severity and predicting suicidality. However, its use depends on accurate automatic speech recognition (ASR). Current gold standard open source ASR systems are trained mainly on adult speech and perform poorly in children, limiting application in pediatric psychiatry.

**Methods:** We benchmarked two open-source ASR models—NVIDIA Parakeet and Whisper-small—on the Ohio Child Speech Corpus (303 children, ages 4–9), using the reference human transcripts provided with the dataset. Audio was standardized to each model’s expected sampling rate. No model fine-tuning or adaptation was performed. For each utterance, we computed word error rate (WER) and character error rate (CER), and assessed semantic fidelity using Sentence Mover’s Distance (SMD) and BERTScore F1. Metrics were summarized overall, stratified by single-year age bins (4, 5, 6, 7, 8, 9), and also grouped into two broader categories: younger children (ages 4–6) and older children (ages 7–9). We compared WER, CER, SMD, and BERTScore F1 across both age groups and evaluated age effects as trends using nonparametric statistical tests.

**Results:** Both models showed significant age effects where younger children had markedly higher word error rates (WER >40%) and character error rates (CER >30%) compared to older children (WER ∼30%, CER ∼20%). Sentence mover distance improved with age, while BERTScore F1 remained stable. Despite age-related improvements, overall transcription accuracy was low.

**Discussion:** Current commonly used open-source ASR systems are inadequate for pediatric audio transcription, specifically in younger children. In order to build clinically translatable tools, collecting child-specific data and model fine-tuning through structured speech paradigms is essential.

## Introduction

### Audio Biomarkers for Psychiatric Disorders

Psychiatric assessments are primarily based on patient subjective reports, clinician observations during the mental status examination that integrate both objective signs and subjective impressions, and the use of standardized clinician-rated and patient self-reported scales to quantify symptom severity and possibly establish a diagnosis or guide possible changes in diagnosis over the course of illness (1). Despite advances in neuroscience and digital health, psychiatric assessments still lack objective technological tools for diagnosis and continue to rely on traditional, subjective methods (2).

Voice biomarkers for mental health remain an underexplored field with a great potential for diagnostic differentiation. For instance, a cross-sectional study observing voice biomarkers in adults with major depressive disorder (MDD), demonstrated that a Machine Learning, Support Vector Machine (SVM) Classifier trained on acoustic and linguistic features such as fundamental frequency, jitter, shimmer, pause structure, and prosodic variability was able to distinguish MDD from healthy controls with an area under curve (AUC) of 0.93(3). However, differentiating between overlapping psychiatric conditions (e.g., Unipolar Depression vs Bipolar Depression) remains a challenge as compared to differentiating against healthy controls. Cross-diagnostic classification studies often achieve only modest accuracies in the 60–70% range, mainly due to limited datasets, and scarcity of standardized speech paradigm protocols(4).

### Current transcription models and lack of pediatric models

Psychiatric audio/speech biomarker studies, including those exploring depression, bipolar disorder, and suicide risk, are dependent on the quality of the underlying transcription models used to process speech recordings. These models belong to the broader class of automatic speech recognition (ASR) systems, which map acoustic waveforms into text representations using deep learning architectures such as recurrent neural networks, transformers, or hybrid approaches(5). Modern foundation-scale ASR systems trained on thousands of hours of speech have achieved impressive robustness to noise, accents, and domain shifts, and can generalize to unseen tasks through zero-shot inference(6). Despite these advances, ASR systems are mainly trained on adult speech, and their performance drops substantially in children(7). Younger children in particular present challenges due to higher fundamental frequency, shorter vocal tract length, immature articulation, variable pronunciation, and frequent disfluencies(8). A key solution to this technological gap is fine-tuning models with child-specific data, specifically data from healthy children and children with psychiatric conditions. That is, using this data to train the models, so that their accuracy becomes sufficient for a clinically translatable tool for differential diagnostic decision-making. Therefore, to establish ASR model accuracy we benchmarked two models - NVIDIA Parakeet and Whisper small model against professionally done human transcription benchmarks. In both models, we evaluated transcription accuracy using the Ohio Child Speech Corpus (OCSC)(9), an open-access dataset of 303 children aged 4–9 recorded in a public museum lab setting. OCSC offers clean lapel-mic audio, diverse structured and spontaneous tasks (e.g., letter-naming, story descriptions, Wug test), and detailed metadata on age, sex, and socio-environmental context. Data were acquired through the Talkbank speech data repository(10). We selected OCSC due to its high-quality child-directed audio, rich annotations, and public availability—making it ideally suited for benchmarking pediatric ASR models and guiding future adaptation for clinical use.

## Methods

Audio data from the Ohio Child Speech Corpus was acquired from talkbank.org and subsequently processed using a custom Python pipeline that leverages CHAT/CHILDES transcript files (.cha)(11) to perform speaker-aware ASR evaluation. The pipeline parsed .cha files using the pylangacq library(11) to extract utterance-level transcripts, speaker identities, and precise timing information (start/end timestamps in milliseconds). For each utterance, the corresponding audio segment was extracted from MP3 recordings using pydub(12), resampled to 16kHz mono, and submitted individually to one of two ASR engines, NVIDIA Parakeet or Whisper. This utterance-level approach eliminated the need for automatic speaker diarization, as ground-truth speaker labels and timing were directly available from the .cha metadata.

We benchmarked **NVIDIA Parakeet**-**TDT**-**0.6B (v2)** and **OpenAI Whisper**-**small** - both compact yet capable models selected for their recent design and manageable inference footprint. Parakeet-TDT-0.6B is a ∼600 million-parameter FastConformer-TDT model (released May 2025) downloadable from Hugging Face. The Parakeet model uses a transducer-style decoder that jointly models tokens and their durations, allowing efficient skipping of silent frames. On the other hand, whisper-small (∼244 million parameters) is a Transformer encoder–decoder model made available through the OpenAI/Hugging Face repository and was trained on ∼680,000 hours of (weakly supervised) speech with strong generalization across domains. Both models report **word error rates in the single-digit (∼ 6-9 %)** based on standard adult English benchmarks as highlighted by the huggingface Opean ASR leaderboad (13) Noticeably, as both models are small, they offer lower latency and memory use - important for real-time or edge deployment - while still achieving competitive WER. Our experiments ran on a single **48 GB NVIDIA RTX 6000 Ada** GPU.

Statistical analyses were performed using traditional metrics including Word Error Rate (WER), Character Error Rate (CER), using the jiwer library(14). Semantic similarity metrics BERTScore(15) F1 and Sentence Mover’s Distance - were calculated using the Bertscore(15) and the sentence transformers library(16) respectively. Statistical analyses and visualization were performed using Python with *scipy stats, seaborn, and matplotlib* libraries. Participant-level ASR metrics were compared across age groups using one-way ANOVA with Tukey’s HSD post-hoc tests, independent samples t-tests for young (ages 4-6) vs. old (ages 7-9) comparisons, and correlation analyses with effect size calculations (Cohen’s d).

## Results

### Dataset Characteristics

The dataset consisted of 303 participants, with an average audio duration of 31.2 minutes per child (range: 9.2–73.5 minutes). Younger children (ages 4–6; n = 139) and older children (ages 7–9; n = 164) contributed to the analyses.

### Nvidia Parakeet

#### Group-Level Comparisons: Younger vs. Older Children

Significant differences in transcription performance were observed across the two age groups. Word Error Rate (WER) was higher in the younger group (mean = 46.5%, SD = 16.9) compared to the older group (mean = 32.2%, SD = 14.4), t(301) = 7.87, p < 0.0001, with a large effect size (Cohen’s d = 0.92). Character Error Rate (CER) followed the same pattern, with younger children showing a mean of 31.3% (SD = 12.4) compared to 21.5% (SD = 10.9) in older children, t(301) = 7.26, p < 0.0001, Cohen’s d = 0.85.

For semantic similarity metrics, Sentence Mover’s Distance (SMD) was significantly lower in older children (mean = 0.15, SD = 0.07) relative to younger children (mean = 0.18, SD = 0.07), t(301) = 3.38, p = 0.0008, Cohen’s d = 0.39. In contrast, BERTScore F1 did not differ significantly between groups, with values of –0.05 (SD = 0.12) for younger children and –0.04 (SD = 0.12) for older children, t(301) = –1.10, p = 0.27, Cohen’s d = –0.13. Utterance count was also comparable between groups, averaging 837 (SD = 261) in the younger group and 876 (SD = 269) in the older group, t(301) = –1.26, p = 0.21, Cohen’s d = –0.14 (Table 1).

**Table 1.**
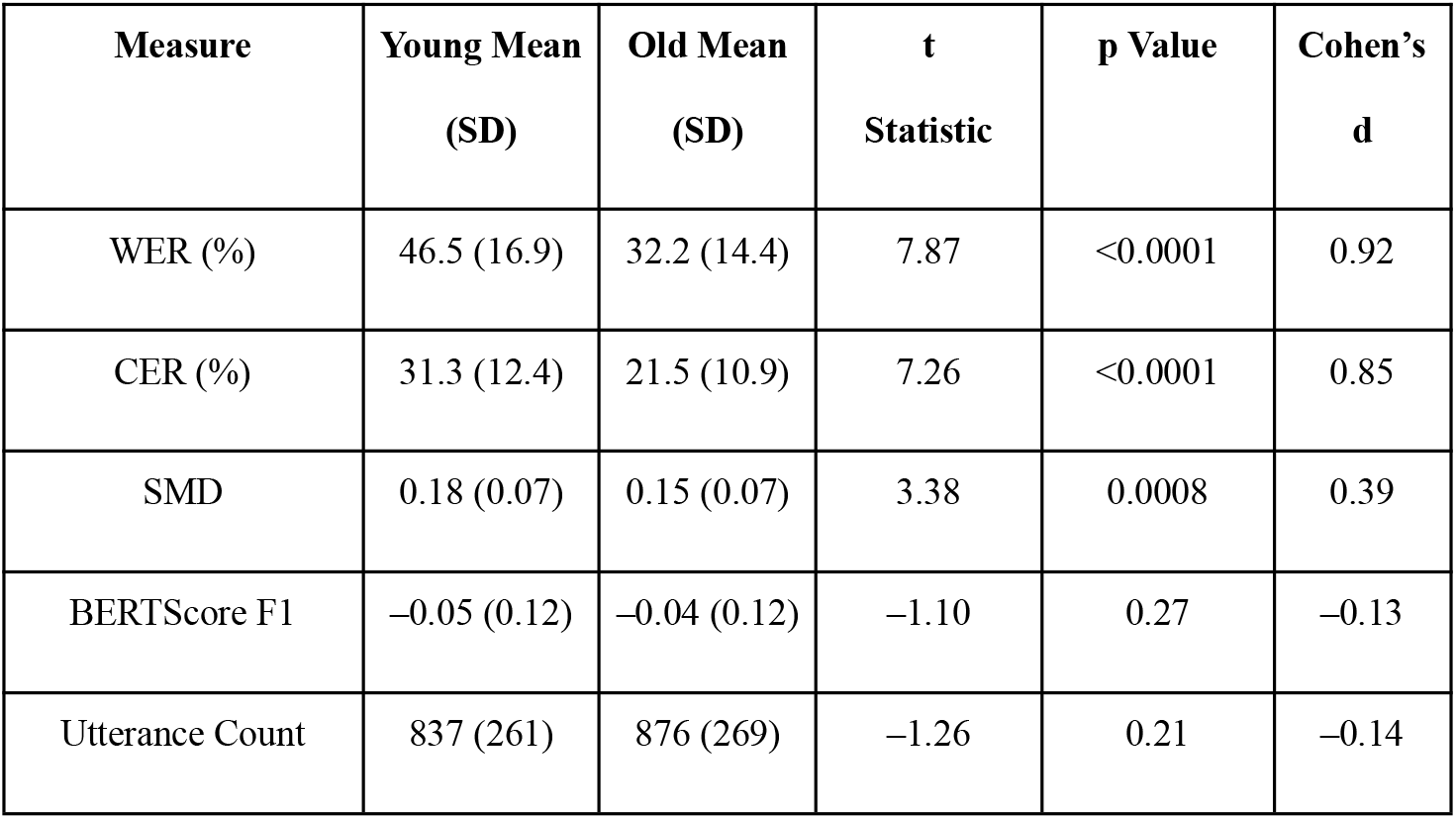
Group comparisons of transcription performance metrics from the Parakeet model between younger and older participants.

| Measure | Young Mean<br>(SD) | Old Mean<br>(SD) | t<br>Statistic | p Value | Cohen's<br>d |
| --- | --- | --- | --- | --- | --- |
| WER (%) | 46.5 (16.9) | 32.2 (14.4) | 7.87 | <0.0001 | 0.92 |
| CER (%) | 31.3 (12.4) | 21.5 (10.9) | 7.26 | <0.0001 | 0.85 |
| SMD | 0.18 (0.07) | 0.15 (0.07) | 3.38 | 0.0008 | 0.39 |
| BERTScore F1 | -0.05 (0.12) | -0.04 (0.12) | -1.10 | 0.27 | -0.13 |
| Utterance Count | 837 (261) | 876 (269) | -1.26 | 0.21 | -0.14 |

#### Age-Specific Effects (4–9 Years)

One-way ANOVAs were conducted to examine transcription performance across individual ages from 4 to 9 years. For WER (Figure 1-A), there was a significant main effect of age (p < 0.0001). Error rates were highest at age 4 (median values above 50%) and progressively declined through ages 5 and 6, reaching the lowest levels at ages 8 and 9 (median values near 25%). Post-hoc comparisons revealed that ages 8 and 9 had significantly lower WER compared to ages 4 through 6 (all p < 0.01).

**Figure 1.**
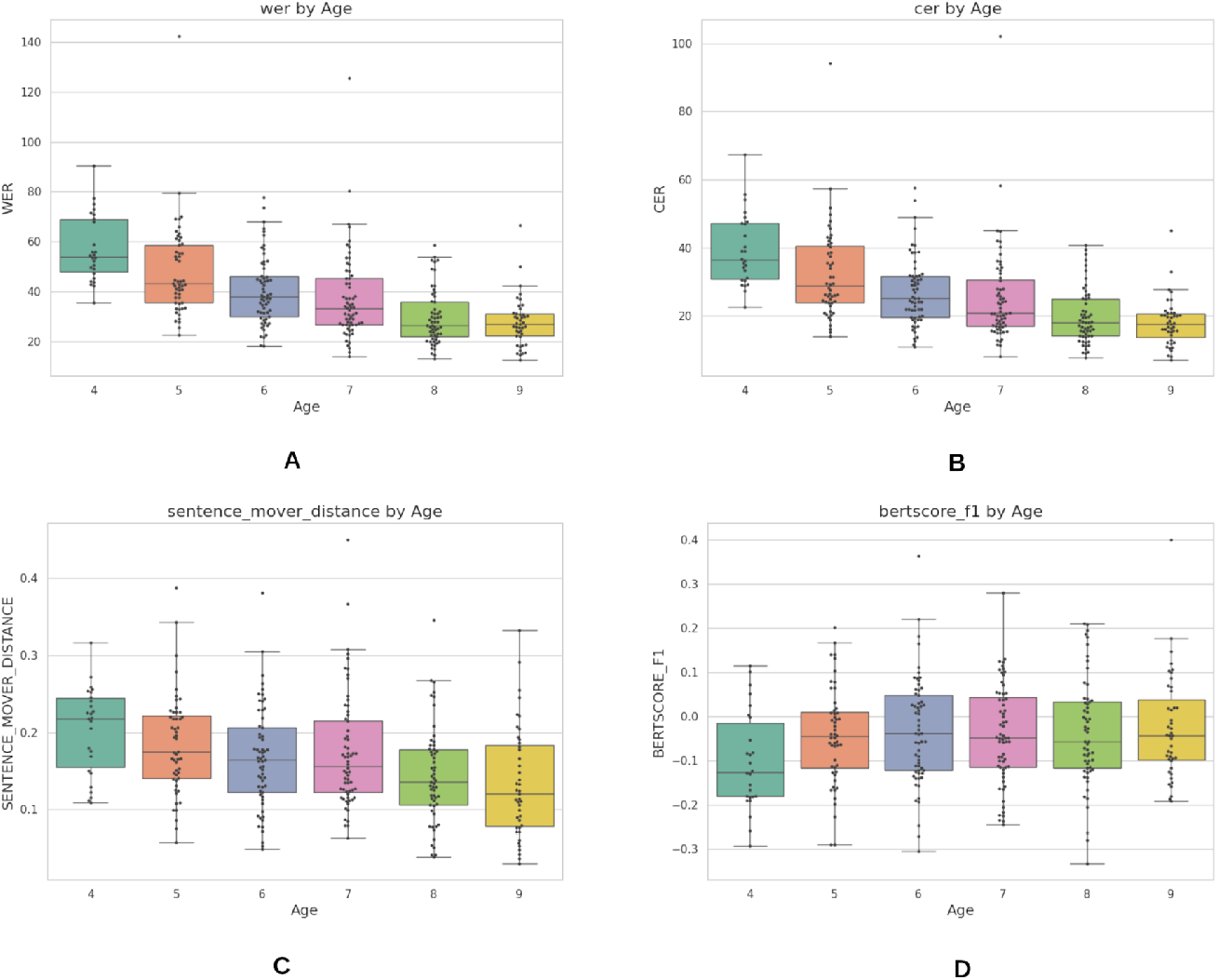
Distribution of automatic speech recognition (ASR) metrics from the Nvidia Parakeet model by age. Each boxplot represents the distribution across participants aged 4–9 years for (A) BERTScore-F1, (B) Character Error Rate (CER), (C) Sentence Mover Distance, and (D) Word Error Rate (WER).

CER (Figure 1-B) also showed a significant effect of age (p < 0.0001). Children aged 4 displayed mean CER values above 35–40%, while those aged 8 and 9 averaged around 15–20%. Post-hoc testing confirmed significant differences between ages 8–9 and ages 4–6, with older children consistently achieving lower error rates.

For semantic similarity, SMD (Figure 1-C) varied significantly across ages (p = 0.0001). Median values were highest in the youngest group (age 4, approximately 0.23) and lowest in the oldest group (age 9, approximately 0.12–0.13). Post-hoc comparisons showed significant reductions between age 4 and ages 8–9, suggesting gradual improvements in semantic fidelity with age.

In contrast, BERTScore F1 (Figure 1-D) did not differ significantly across ages (p = 0.16). Scores remained relatively stable, with median values centered around –0.05 to –0.02 across all ages, and no clear monotonic trend.

#### Correlation Analyses

Correlation analyses (Figure 2) demonstrated strong relationships among performance metrics. WER and CER were highly correlated (r > 0.8, p < 0.001), confirming that both measures captured overlapping variance in transcription accuracy. Both WER and CER were negatively correlated with BERTScore F1, indicating that lower error rates were associated with improved embedding-based similarity. Conversely, WER and CER were positively correlated with SMD, showing that higher error rates corresponded to greater semantic distortion in the transcriptions. These associations underscore the consistency across error-based and semantic measures.

**Figure 2.**
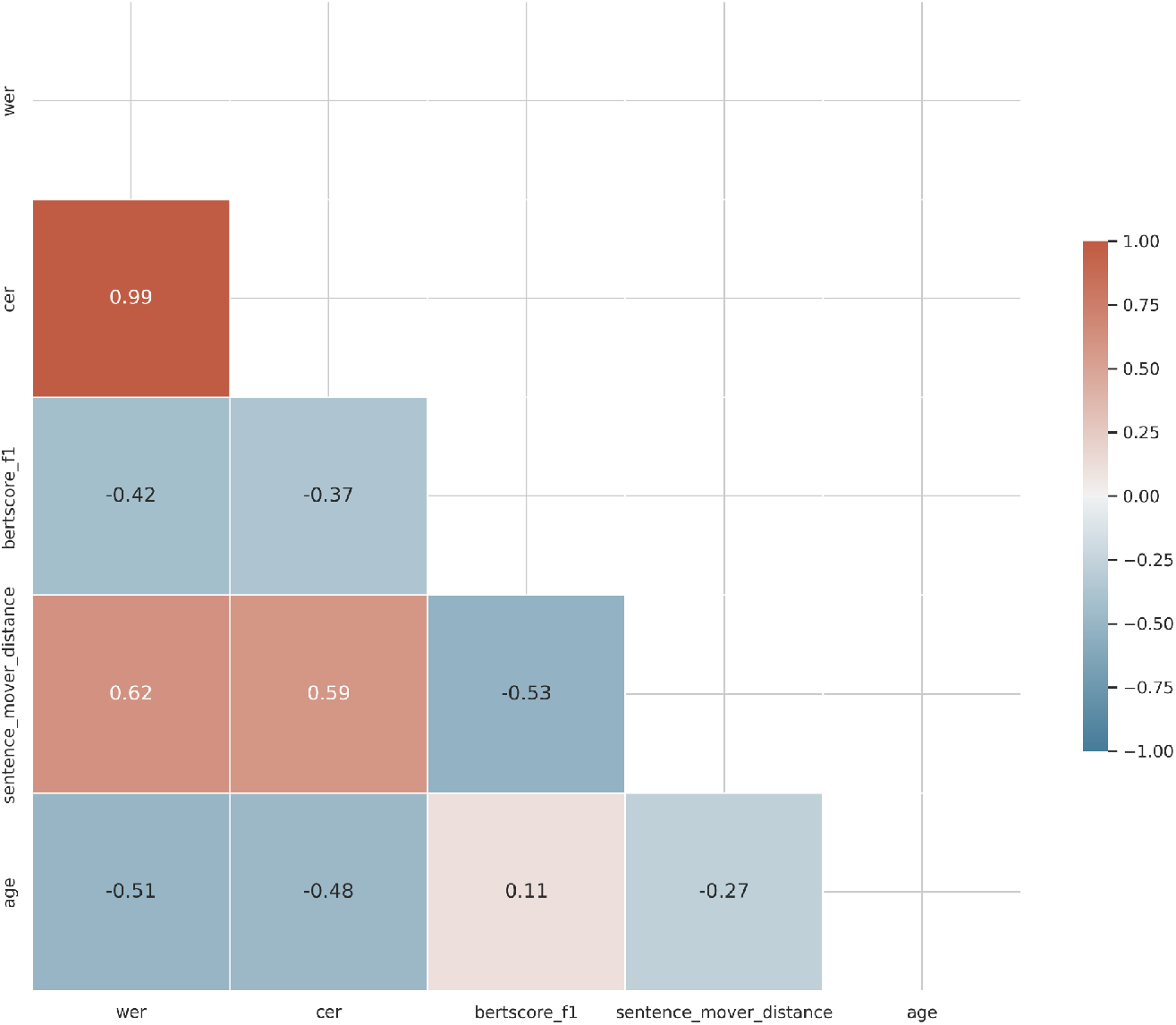
Correlation matrix of Nvidia Parakeet model performance metrics and age. Positive correlations are shown in red and negative correlations in blue.

### Results – OpenAI Whisper Small Model

#### Group-Level Comparisons: Younger vs. Older Children

Significant differences in transcription performance were observed across the two age groups. Word Error Rate (WER) was higher in the younger group (mean = 41.3%, SD = 15.1) compared to the older group (mean = 29.0%, SD = 12.7), *t*(301) = 7.58, *p* < 0.0001, with a large effect size (Cohen’s *d* = 0.89). Character Error Rate (CER) followed the same pattern, with younger children showing a mean of 29.8% (SD = 16.5) compared to 20.3% (SD = 10.0) in older children, *t*(301) = 5.91, *p* < 0.0001, Cohen’s *d* = 0.71.

For semantic similarity, Sentence Mover’s Distance (SMD) was significantly lower in older children (mean = 0.14, SD = 0.07) relative to younger children (mean = 0.16, SD = 0.06), *t*(301) = 2.61, *p* = 0.0096, Cohen’s *d* = 0.30. In contrast, BERTScore F1 did not differ significantly between groups, with values of –0.0004 (SD = 0.12) for younger children and –0.0036 (SD = 0.11) for older children, *t*(301) = 0.24, *p* = 0.81, Cohen’s *d* = 0.03. Utterance count was also comparable between groups, averaging 823 (SD = 228) in the younger group and 847 (SD = 201) in the older group, *t*(301) = –0.95, *p* = 0.34, Cohen’s *d* = –0.11 (Table 2).

**Table 2.** Group comparisons of transcription performance metrics from the OpenAI Whisper Small model between younger and older participants.

| Measure | Young Mean<br>(SD) | Old Mean<br>(SD) | t Statistic | p Value | Cohen's d |
| --- | --- | --- | --- | --- | --- |
| WER (%) | 41.3 (15.1) | 29.0 (12.7) | 7.58 | <0.0001 | 0.89 |
| CER (%) | 29.8 (16.5) | 20.3 (10.0) | 5.91 | <0.0001 | 0.71 |
| SMD | 0.16 (0.06) | 0.14 (0.07) | 2.61 | 0.0096 | 0.30 |
| BERTScore<br>F1 | −0.0004<br>(0.12) | −0.0036<br>(0.11) | 0.24 | 0.81 | 0.03 |
| Utterance<br>Count | 823 (228) | 847 (201) | −0.95 | 0.34 | −0.11 |

#### Age-Specific Effects (4–9 Years)

One-way ANOVAs were conducted to examine transcription performance across individual ages from 4 to 9 years.

For WER (Figure 3-A), there was a significant main effect of age (*p* < 0.0001). Error rates were highest at age 4 (median values >50%) and progressively declined through ages 5 and 6, reaching the lowest levels at ages 8 and 9 (median values near 25%). Post-hoc comparisons revealed that ages 8 and 9 had significantly lower WER compared to ages 4 through 6 (all *p* < 0.01).

**Figure 3.**
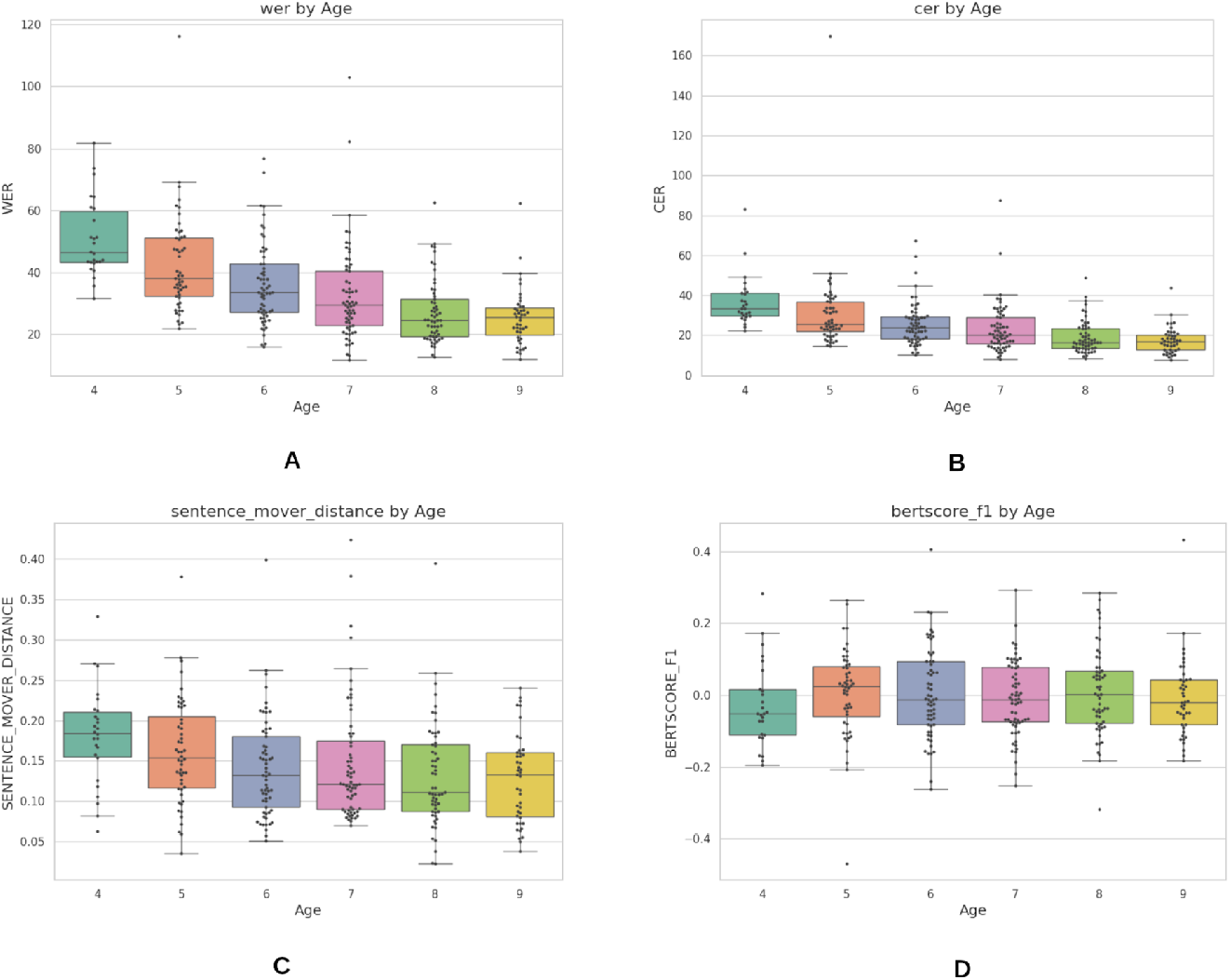
Distribution of automatic speech recognition (ASR) metrics from the OpenAI Whisper Small model by age. Each boxplot represents the distribution across participants aged 4–9 years for (A) BERTScore-F1, (B) Character Error Rate (CER), (C) Sentence Mover Distance, and (D) Word Error Rate (WER).

CER (Figure 3-B) also showed a significant effect of age (*p* < 0.0001). Children aged 4 displayed mean CER values above 35–40%, while those aged 8 and 9 averaged around 15–20%. Post-hoc testing confirmed significant differences between ages 8–9 and ages 4–6, with older children consistently achieving lower error rates.

For semantic similarity (Figure 3-C), SMD varied significantly across ages (*p* < 0.0001). Median values were highest in the youngest group (age 4, approximately 0.22–0.23) and lowest in the oldest group (age 9, approximately 0.12–0.13). Post-hoc comparisons showed significant reductions between age 4 and ages 8–9, suggesting gradual improvements in semantic fidelity with age.

In contrast, BERTScore F1 (Figure 3-D) did not differ significantly across ages (*p* = 0.16). Scores remained relatively stable, with median values centered around –0.05 to –0.02 across all ages, and no clear monotonic trend.

#### Correlation Analyses

Correlation analyses (Figure 4) demonstrated strong relationships among performance metrics. WER and CER were highly correlated (r > 0.98, *p* < 0.001), confirming that both measures captured overlapping variance in transcription accuracy. Both WER and CER were negatively correlated with BERTScore F1, indicating that lower error rates were associated with improved embedding-based similarity. Conversely, both error metrics were positively correlated with SMD, showing that higher error rates corresponded to greater semantic distortion. Age was negatively correlated with both WER and CER, further confirming that transcription accuracy improved with development.

**Figure 4.**
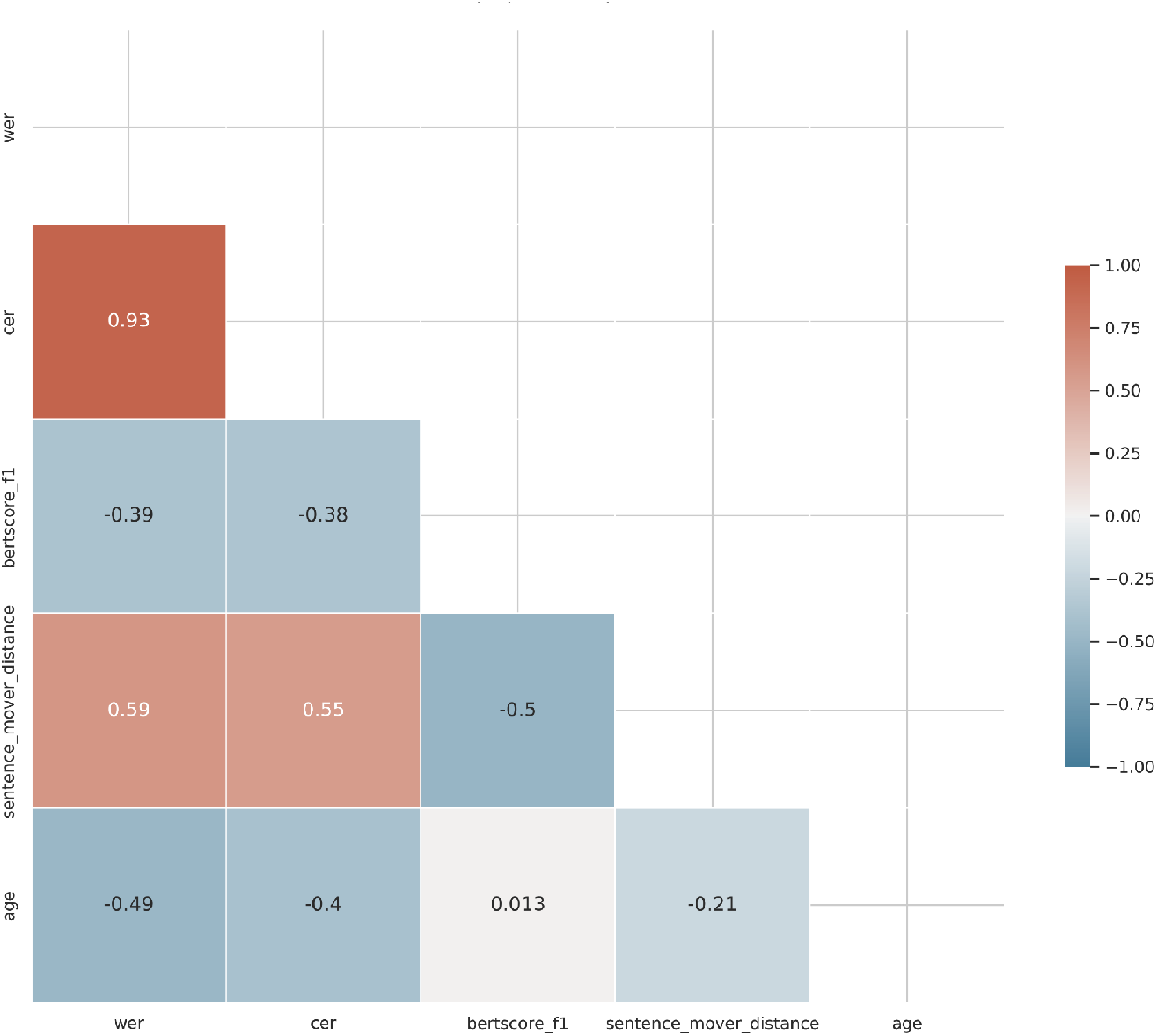
Correlation matrix of OpenAI Whisper Small model performance metrics and age. Positive correlations are shown in red and negative correlations in blue.

## Discussion

The NVIDIA Parakeet model showed large developmental effects but overall high transcription error rates across the pediatric sample. In group-level comparisons, WER averaged 46.5% (SD = 16.9) in younger children (ages 4–6) and 32.2% (SD = 14.4) in older children (ages 7–9). CER values followed the same pattern, decreasing from 31.3% (SD = 12.4) in the younger group to 21.5% (SD = 10.9) in the older group. Semantic fidelity, indexed by Sentence Mover’s Distance, improved modestly with age (0.18 vs. 0.15). However, these error rates remain high compared to adult benchmarks, and BERTScore F1 did not show meaningful differences across groups. ANOVA confirmed significant main effects of age for WER, CER, and SMD, with the steepest contrasts between ages 8–9 and 4–6. Correlations confirmed that higher error rates were tightly coupled with lower semantic fidelity.

The Whisper Small model revealed a nearly identical developmental trajectory, with even younger children experiencing error rates exceeding 40% WER and 30% CER. Older children performed somewhat better, with WER values in the low 30% range and CER around the low 20s, but performance was still far below levels required for reliable clinical transcription. SMD decreased with age, indicating improved semantic similarity, but absolute values remained poor compared to adult-level ASR. As with Parakeet, BERTScore F1 did not differ across groups, and utterance counts were stable. Age-stratified analyses showed significant declines in error measures from age 4 to ages 8–9, but the absolute performance at all ages was far worse than typical adult speech recognition.

Taken together, these findings highlight the current technological limitation: neither Parakeet nor Whisper Small provide adequate transcription performance for pediatric speech, particularly for younger children. Both models demonstrate systematic age effects, but even at ages 8–9, error rates remain too high for dependable downstream use in psychiatric audio biomarker research. The extremely poor performance in the youngest children underscores that current open-source ASR systems are not trained for child speech and cannot yet serve as a foundation for pediatric psychiatry applications without substantial adaptation or fine-tuning. These results collectively point to a clear gap in the field: there are currently no robust, validated open-source transcription models for pediatric psychiatry, and meaningful clinical progress will require dedicated pediatric data and fine-tuning approaches.

The development of clinically translatable pediatric ASR systems would open pathways to the development of translational differential diagnostic and monitoring tools in psychiatry. Voice features have already been linked to symptom severity and treatment response in adult depression and bipolar disorder(17), and extending these approaches to pediatric populations could support earlier identification and longitudinal tracking. A particularly promising avenue is distinguishing pediatric unipolar versus bipolar depression(18), a clinically challenging differential diagnosis where early, objective tools may improve treatment allocation. Beyond mood disorders, standardized voice tasks may help differentiate autism spectrum disorder (ASD) and ADHD(19), or aid in parsing overlapping neurodevelopmental and psychiatric conditions.

## Data Availability

All data produced in the present work are contained in the manuscript

## Acknowledgments

This work was supported by the NECMHR01 grant from the Texas Child Mental Health Care Consortium (TCMHCC) and the Dunn Foundation. Speech dataset acquired from the Ohio Child Speech Corpus (OCSC) through talkbank.org

